# Organizational culture, quality of care and leadership style in government general hospitals in Kuwait: a multimethod study

**DOI:** 10.1101/2020.07.15.20154245

**Authors:** Talal ALFadhalah, Hossam Elamir

## Abstract

**Purpose:** To investigate the organizational culture, assess the quality of care, and measure their association with a transformational/transactional leadership style in six hospitals.

**Materials and methods:** We used cross-sectional and retrospective quantitative approaches in government-sponsored secondary-care hospitals. A sample of 1,626 was drawn from a frame of 9,863 healthcare workers in six hospitals. Followers were surveyed using the Multifactor Leadership Questionnaire and the Organizational Description Questionnaire. We reviewed and analyzed one year (2012) of quarterly and annual quality indicators from the hospitals. Data were analyzed using suitable statistical analyses.

**Results:** We collected 1,626 responses from six hospitals. 66.4% to 87.1% of participants in each hospital identified their hospital’s organizational culture as transformational, whereas 41 out of 48 departments were identified as having a transformational culture. The percentage of participants at each hospital rating their leader and organizational culture as transformational ranged from 60.5% to 80.4%. The differences between leadership style and organizational culture were statistically significant for four of the hospitals. For most of the quality indicators, there was a positive, but nonsignificant, correlation with leadership style.

**Conclusion:** Leaders define and influence organizational culture. The prevailing transformational leadership style creates and maintains a transformational organizational culture. The effect of transformational leadership on the quality of care delivered by the organization was measured in this study, and showed a positive and nonsignificant relationship between generic quality indicators and the transformational style.

## Introduction

In healthcare organizations, nothing plays a more significant role in shaping organizational culture, improving quality of care, and enhancing patient safety than leadership.^1–3^ Leadership has been described as “*a process whereby an individual influences a group of individuals to achieve a common goal”*.^4(p6)^ Since the emergence of a definition that emphasizes control, domination, and centralization of power, the topic of leadership has attracted a sizable number of researchers.^4,5^ For over more than a century, factors such as politics and researchers’ perspectives have influenced the emergence and evolution of leadership theories that include the trait approach, the skills approach, the behavioral approach, the situational approach, path–goal theory, and leader–member exchange theory.^4^ Moral approaches, such as ethical and authentic leadership, have gained traction since the start of the new millennium.^4,6^

Healthcare executives, directors, and managers can enact different leadership styles and influence their followers’ actions.^3,7–9^ Leaders approximate or choose their style based on a combination of their beliefs, values and performance, with contributions from organizational culture and norms, which favor some leadership styles and disfavor others.^3,10^ The most commonly researched and applied leadership theories are those founded on relational aspects of leadership, particularly transformational leadership.^5,11,12^ Transformational leadership is “*the process whereby a person engages with others and creates a connection that raises the level of motivation and morality in both the leader and the follower*”.^4(p186)^

Transformational leaders can help to develop, shape, and maintain a desired organizational culture. They do so by creating and infusing the values, beliefs, and perceptions that they believe are necessary and good for the organization.^3,13^ Manifested in some characteristics as organizational innovation and learning, organizational culture is defined as the shared basic assumptions learned by staff that distinguish their organization from other similar entities.^6,14^ Organizational culture is a variable that significantly influences an organization’s outcomes. A leader’s interactions with followers, their approach to addressing problems, reaction to competition, and implementation of new strategies, all influence organizational culture.^3,15^ The stronger and more unified the staff values, beliefs, and perceptions are, the stronger the organizational culture.^6,14^ A predominant culture in a healthcare organization ensures consistent behavior between its members, which reduces conflict and creates a healthy working environment.^3,16^

Quality of care is an indispensable component of a healthcare organization’s performance.^13,17^ It depends on many factors, such as planning and provision of services that meet patients’ needs, acquiring and allocating resources, providing sufficient staff, nurturing a culture that fosters quality and safety, and setting priorities for improvement.^7,13,18^ The World Health Organization characterizes high-quality healthcare services as effective, safe, and people-centered.^19^ Only healthcare leaders have the resources and control to exhibit characteristics that influence and support good quality and safety.^3,18,20^ Healthcare quality can be assessed by quality indicators, quantitative measures used to evaluate and monitor the processes of care, customer service, and different aspects of the organization that are known to contribute to the quality of its outcomes.^21^ Generic quality indicators are one type that measure aspects of care relevant to most patients regardless of their diagnosis or care setting.^22,23^

Acknowledging that this field is under-researched in the Kuwait/Arab region despite its high importance and impact,^21,24^ the objective of this paper is to explore and assess organizational culture and quality of care, and measure their association with transformational/transactional styles of leadership in the hospitals studied. The rationale behind focusing on transformational leadership is because it is among the most recurrent theories in research,^5,11^ in addition to being one of the most effective leadership styles in health services,^25,26^ and has a prominent impact on growth of leadership development strategies.^12^ This paper is the second from a research project aimed at assessing leadership styles, organizational culture, patient safety initiatives, and quality of care in six government general hospitals in Kuwait. The first paper reported the leadership styles in the six government general hospitals.^24^ We present here an analysis and discussion built on the previous finding that transformational leadership is predominant.

## Material and methods

### Setting

This was a multicenter study conducted at the six government general hospitals (coded A, B, C, D, E, and F) in Kuwait. At the time of data collection, the government healthcare system in Kuwait was providing the majority of secondary healthcare services at these six general acute care hospitals.^24^ Hospital beds ranged between 398 and 866 in number.^27^ The six hospitals have the following clinical and allied health departments: medicine, surgery, pediatrics, intensive care unit (ICU), accident and emergency (A&E), laboratory, nursing, and pharmacy.

### Study instrument and data collection

This is a multimethod study conducted with cross-sectional and retrospective quantitative approaches. A period of one month in 2013 was spent in each hospital collecting data from followers using two self-administered paper-based questionnaires: the Multifactor Leadership Questionnaire (MLQ) and the Organizational Description Questionnaire (ODQ).^28,29^ These questionnaires have two versions, one to be answered by the leader, the other by followers. In this paper, survey respondents are referred to as followers.

The heads of the quality offices were assigned as points of contact in their respective hospitals. They approached potential participants in their break rooms and explained the aims and requirements of the study. Those who voluntarily agreed to participate received copies of the study instrument inside envelopes marked with a unique identifier code. Participants’ names were recorded under this code in a register to facilitate the retrieval of answered questionnaires inside the sealed envelopes. The registers were collected and retained by the lead author, who destroyed them upon receiving all the completed forms.

The MLQ consists of 45 items that cover different factors of leadership: characteristic of transformational leadership, characteristic of transactional leadership, non-transactional (laissez-faire) leadership, and the outcome of leadership. For example, “Avoids making decisions” is a sample item in the “Rater Form”, which the respondent rates on a five-point scale: 0 = Not at all, 1 = Once in a while, 2 = Sometimes, 3 = Fairly often, and 4 = Frequently, if not always.^29^

The ODQ consists of 28 statements split into two: 14 odd-numbered statements support a profile of transactions, and 14 even-numbered statements deal with transformational attributes. Each statement describes the general organizational behavior and beliefs of transformational or transactional leaders. The respondents were asked to indicate whether they believe the statement is true (T) or false (F) for their organization. A third category (?) can be selected if the respondent is indecisive or cannot say. “We negotiate with each other for resources” is a sample transactional statement, whereas “We trust each other to do what’s right” is a sample transformational statement.^28^

The retrospective quantitative approach reviewed and included statistics on the generic quality indicators only. These indicators are five in number and were collected in each hospital for the year 2012.^30^ They are analyzed quarterly and reported annually as a measure of the quality of care in the government hospitals of Kuwait. The indicators were developed by the Quality and Accreditation Directorate in 2002 for use in all government hospitals in Kuwait. They are:

– Percentage of patients discharged against medical advice in inpatient departments.
– Percentage of elective operations cancelled on the day of, or after, admission.
– Percentage of lengths of stay for appendectomy operations of five days and over.
– Percentage of patients discharged from the general surgical department without undergoing an operation.
– Unscheduled return to operating theatre within 48 hours during the same hospital admission.

Although four of the five indicators are surgical, all have a target to decrease the percentage/number. In other words, the lower the indicator result, the better the performance of the hospital and the quality of care. Table 1 presents more information about the five indicators.

### Study population and sampling

We preferred that followers have direct contact with their respective leader, and thus we excluded trainees and assistant registrar physicians and technicians. Subjects who had spent less than one year in the hospital were also excluded. Hence, the study population consisted of 9,863 individuals representing the following professions: physician, nurse, and pharmacist. The population size of professionals in all categories in the six hospitals ranged between 1,448 and 1,961. Based on a previous study on leadership styles,^31^ the required sample size for this study was calculated using STATA 10 to be 271 per hospital using STATA 10. The calculation was performed assuming:

1. A mean score of employees’ perception of their leader as a transformer = 24.62.
2. A standard deviation (SD) of = 8.81 and accepted error = 1.5.
3. An alpha of 0.05 and a power of 80%.^32^

Physicians in each hospital from different departments, nurses, and pharmacists were selected using proportional allocation. We aimed to keep the sample size from each stratum proportional to the stratum size. The proportional allocation provides a self-weighted sample and requires no additional weighting to estimate unbiased population parameters.^33,34^ As the rotation schedules of nurses typically differ by department, the sample of nurses was randomly selected in each hospital from all departments.

### Data management and analysis

Transactional and transformational leadership style and organizational culture scores were calculated from the MLQ and the ODQ data, respectively, as follows:^28,29^

1. MLQ: Items on the MLQ are rated on a five-point scale. We used Formulae 1 and 2 to calculate the mean scores of leadership styles:

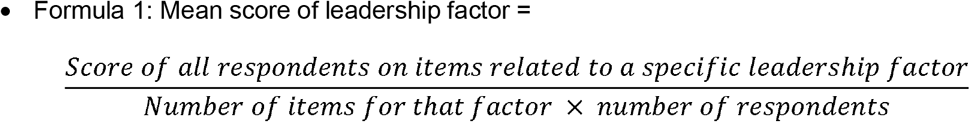

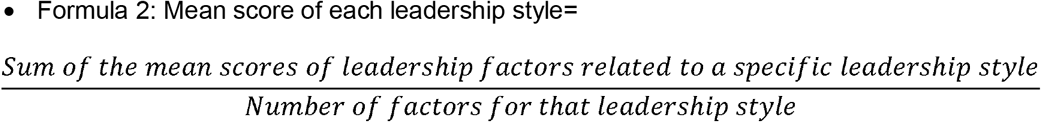
  - The higher of the two scores from Formula 2 indicated whether leadership was transactional or transformational.
2. ODQ:
  - Transactional culture score: add one for each odd-numbered statement marked as true and subtract one for each odd-numbered statement marked as false.

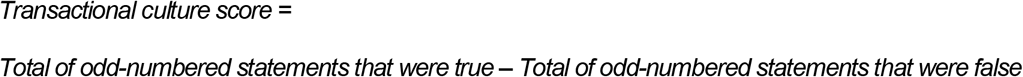
  - Transformational culture score: add one for each even-numbered statement marked as true and subtract one for each even-numbered statement marked as false.

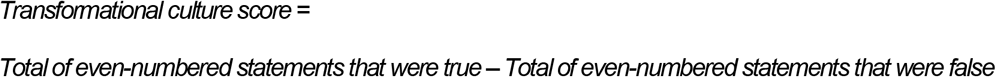
  - The indecisive or cannot say statements (ã) were scored as zero.
  - The higher of the two scores indicated whether the culture was transactional or transformational.

SPSS 23.0 was used to analyze the data. A *p* value ≤ .05 was considered to indicate statistical significance. The analysis of the quantitative data included univariate descriptive (means, standard deviations, frequencies, percentages) and bivariate (chi-squared tests, Pearson’s correlation) analyses to examine the association between the leadership style, organizational culture, and quality of care.

### Ethical approval and consent to participate

Ethical approval and consent to conduct the study was granted by the Standing Committee for Coordination of Health and Medical Research in Kuwait (219/2012). We confirm that all methods were conducted in accordance with the relevant guidelines and regulations of the Standing Committee for Coordination of Health and Medical Research in Kuwait. Participating hospitals provided permission for the study to take place, and hospital and respondent identities were coded to ensure anonymity and remained confidential. Participants provided voluntary verbal informed consent after receiving an explanation of the value, benefits, and risks of the study, and their questions were satisfactorily answered. Verbal informed consent was approved by the Standing Committee for Coordination of Health and Medical Research in Kuwait.

## Results

With 1,626 responses from six hospitals, the response rate for the two questionnaires was 100%. Table 2 shows the demographic and work-related characteristics of followers in these hospitals. The age group 30–39 years was the most represented age group in all six hospitals, followed by 40–49 years. Females represent two-thirds of the sample, and non-Kuwaitis represent seven-eighths. In hospitals B and D, the relative majority held a graduate degree, whereas in hospitals E and F the relative majority held a diploma. Other than age and category of pharmacist, all the followers’ demographic and work-related characteristics show statistically significant differences between the six hospitals.

The complete analysis of the MLQ is outside the scope of this paper and was reported previously.^24^ It showed that *“all followers rated their leaders as transformational leaders, except the followers of the head of pharmacy in hospital A and the head of surgery in hospital C, who rated their leaders as transactional”*.^24(p463)^ Notably, followers rated their transformational leaders with lower scores than the leaders’ self-rating, except for the heads of medicine in hospitals A and E. The same was true of heads of ICU, A&E, and Laboratory in hospitals B, E, and C, respectively. Another relevant finding is the low mean scores (2.35–2.86) of heads of surgical departments. Among all heads of departments, heads of surgical departments achieved the lowest ratings as transformational leaders in three hospitals (D, E, and F) and the second lowest in two hospitals (A and B). The head of surgery in hospital C was identified as a transactional leader. Appendix 1 shows the data relevant to this part of the study.

A comparison of the organizational cultures of the six hospitals shows that hospital D had the highest percentage rating for a transformational style (87.1%), followed by hospitals C and F (80.1% and 75.3%, respectively) (Table 3). The lowest rating was recorded for hospital B (66.4%). Departmental organizational culture styles were mainly transformational except in Surgery, ICU, and Pharmacy in hospital E (52.9%, 55.6%, and 75.0%, respectively), A&E and Laboratory in hospital D (60.0% and 66.7%), and Medicine in hospital B (59.1%), where it was a transactional culture. In hospital C, transformational and transactional cultures were equally represented in the pharmacy department (50% each). Across the hospitals, the departments of Medicine and Nursing showed statistically significant differences in regards to organizational culture. Two hospitals (D and E) showed statistically significant differences with respect to organizational culture across the departments.

Table 4 indicates that the percentage of followers who believed that they have both a transformational leader and transformational organizational culture ranged from 60.5% (hospital B) to 80.4% (hospital D). The percentage of followers who believed that they have a transactional leader, as well as a transactional organizational culture, ranged from 4.1% (hospital E) to 7.7% (hospitals A and B). The percentage of followers who believed that they have a transformational leader, but rated their organizational culture as transactional ranged from 8.5% (hospital D) to 25.8% (hospital B). The differences between leadership style and organizational culture were statistically significant for hospitals A, B, E, and F, with *p* values of ≤.001, ≤.001, .002, and .002, respectively.

Table 4 also shows the generic quality indicator (four surgical, one nonsurgical) results from 2012 in the six hospitals. Hospital F had the highest percentage of discharges against medical advice (7.9%) and hospital B had the lowest (1%). Hospital B had the highest percentage of cancelled operations (12.9%) and hospital D had the lowest (9.3%). Hospital C had the highest percentage of length of stay of five days or longer after appendectomy (46.6%) and hospital D had the lowest (14.1%). Hospital D had the highest percentage of discharge from the surgical department without an operation (54.5%) and hospital E had the lowest (32.3%). The differences between hospitals with respect to these four indicators are statistically significant. For the number of unscheduled returns for operations, hospital A had the highest (56 cases) and hospital C had the lowest (9 cases).

Table 5 shows the relationship between generic quality indicators and the transformational leadership style of heads of departments in the six hospitals. It is worth noting that a negative correlation is desired. The correlation between transformational leadership style and both the percentage of discharge against medical advice and percentage for a length of stay for five days or longer after appendectomy were very weak,^35^ nearly null, negative, and nonsignificant (*r* = −0.03, *p* = .957 and *r* = −0.09, *p* = .872, respectively). The correlation between transformational leadership and the percentage of cancelled operations was moderate,^35^ negative, and nonsignificant (*r* = −0.37, *p* = .468). The correlation between transformational leadership and the number of unscheduled return for operations was strong,^35^ negative and nonsignificant (*r* = −0.71, *p* = .111). The correlation between a transformational leadership style and the percentage of discharge from the surgical department without operation was moderately^35^ positive, but nonsignificant (*r* = 0.49, *p* = .329).

## Discussion

The results of the study reveal that there was a greater frequency of respondents rating their hospital culture more transformational than transactional. These findings could be explained by hospital leaders more often displaying a transformational leadership style than a transactional one. This likely has a great effect on shaping and preserving the culture of the hospital. The results also indicate that the majority of followers that view their department heads as transformational, considered their organizational culture as transformational. This is consistent with the findings of many studies that assessed the relationship between transformational leadership style and transformational organizational culture and reported that there was a significant correlation and positive impact with overall transformational leadership practices.^3^ Moreover, the transformational leadership style has a positive and significant impact on organizational innovation^36,37^ and learning,^38,39^ which are among the primary components of the essence of organizational culture.^14^ This might also explain how the transformational leadership style indirectly creates a transformational culture.

Many studies have found a relationship between transformational leadership style and the quality of care in hospitals.^3,40,41^ Because the generic quality indicators of the government health system report the unwanted occurrences—the lower the better—this study shows that there was a negative nonsignificant correlation between a transformational leadership style and most of the indicators analyzed (Table 5). This relationship might exist because of the support and follow up the transformational leaders provide to their hospital quality officers, who are responsible for the implementation of these generic indicators. Moreover, the characteristics of the transformational leadership style, such as influencing, advising, and being attentive to followers’ needs could be factors in the improvement of the followers’ performance, reflecting an amelioration of the hospital’s quality indicator statistics. That the correlation is statistically insignificant implies the presence of confounding factors that should be investigated.

Many factors might have contributed to the nonsignificance of the mostly weak to moderate correlation between leadership styles and generic quality indicators. Firstly, the percentages on transformational culture within the surgical departments are statistically insignificant (Table 3). Moreover, the mean scores of the transformational heads of surgery are among the lowest compared to other departments (Appendix 1). Given the fact that four of the five reported indicators are surgical, the low scores of the heads of surgical departments largely explain the weak effect transformational leadership has on the quality indicators. Secondly, the studied indicators reflect the performance of the hospital as a whole, whereas leadership styles are assigned to individuals. The third factor is related to the discrepancies between the scores from self-ratings and followers’ ratings, which is associated with a more negative organizational culture. Authors noted that if leaders rated themselves more positively than their followers, hospital performance, in general, might be affected.^42^ To overcome such a limitation, studies recommend to train and educate current and future leaders on the topic of leadership, including its styles and the effective use of its strengths.^1,2,8,41,43^

A recent meta-analysis found that increasing transformational behavior might strengthen any positive impact, however weak, of this trainable leadership style on staff performance.^44^ This analysis is highly relevant to the current study because it addresses two issues: the low scores of transformational leaders and the weak impact of transformational leadership on quality. Being a trainable leadership style, acquiring and improving transformational behavior will be an extremely desirable goal. Once leaders improve their transformational behavior, their impact on improving quality of care will be profound.

### Strengths and limitations

The majority of articles published on the topic of transformational leadership and quality of care assessed the style of leadership in nursing. This study was conducted in multiple centers that represent the country-wide secondary healthcare services, and included a relatively large number and variety of professions and authority levels among the participants. The multimethod design of the study allowed the exploration of several relationships between different components and subjects. It also facilitated triangulation and a wider view of the topic. Furthermore, this is the first study in Kuwait and the region to report on transformational leadership style, transformational organizational culture, quality of care indicators, and their interrelationships. Also, we overcame the potential biases of case series and case reports with the cross-sectional design, which allowed the collection of data for measuring different variables in the population sample at a single point in time.^45^

However, some limitations must be acknowledged. Although a recent systematic review and a research article from the region reported the predominance of transformational leadership,^46,47^ some might claim that the transformational leadership represented in the current study reflects social desirability bias, cultural influences, hiring practices, and management education. Another bias that might have affected our study is selection bias, as we selected quality indicators solely for their availability. The small number of quality-of-care indicators, being primarily surgical, and the nature of reporting unwanted occurrences did not allow the robust evaluation of the quality of care that we sought. In addition, there might be other confounding factors not studied here that resulted in the nonsignificant negative correlation between quality indicators and a transformational style of leadership.

Also, the study could not investigate differences in leadership style based on the nationality of the leaders, because 63 of the 66 leaders studied were Kuwaiti. The study included neither private sector hospitals nor other care delivery settings within the government sector (primary, tertiary, and quaternary healthcare services). Finally, the study explored the relationship between culture and quality with only one leadership model. This does not merit conclusions about the inferiority or superiority of a transformational leadership style over other leadership styles or say anything about how other leadership styles affect culture or outcomes in healthcare organizations.^2,48^

### Practice implications

The requirement for continuing professional development in the healthcare profession makes a culture of learning and transforming more desirable. Therefore, training and development programs are essential for leaders to develop a strong vision and philosophy to communicate expectations, develop others, and lead healthcare organizations to meet strategic objectives.^1,2,41^ This is critical for those in management roles within Kuwait’s health system because they rarely undertake adequate training in related fields.^49^ Fortunately, the transformational leadership style has a noticeable influence on how leadership development strategies evolve.^12^

This study provides insight into a complex and important regionally under-researched area. We invite researchers to explore and compare the different leadership styles and models. We desire further collaboration with the quality indicators team to reflect on how to advance the current indicators program.

## Conclusion

According to followers’ ratings, organizational culture in the six studied hospitals is mostly transformational, with large percentages of followers rating their leader and organizational culture as transformational. In general, our results in this context suggest that leaders are shapers and influencers of their organizational culture.

The transformational leadership style has a positive effect on the quality of care delivered by an organization.^3,40,41^ This effect can be measured using indicators that compare the healthcare organization’s performance to an external reference or “gold standard”. However, it is not enough to be a transformational leader, leaders have to improve their transformational behavior to maximize the gains of this effective leadership style.^43,44^

The relationship between a transformational leadership style and quality indicators was measured in this study. There is a positive impact, of mostly weak to moderate magnitude, of transformational leadership on the quality of care represented by generic quality indicators. However, this impact was found to be statistically insignificant for a couple of possible reasons. The results suggest an opportunity exists to enhance the quality of care if transformational leadership could be improved. Effective transformational leadership can be improved through training, education, experience, and professional development. This field should be further explored to conceptualize the confounding and mediating factors that impact the effectiveness of a practiced leadership style.

## Data Availability

Data are available upon request.

## Acknowledgments

The authors acknowledge and thank Prof. Hoda H. Zaki for her guidance and contribution.

## Notes

This work was supported by the Ministry of Health (MOH) in Kuwait. The MOH (grant number 219/2012) covered the costs of transportation, purchasing the questionnaires from the publisher, writing and printing the data collection tools, and statistical analysis of the data.

## Abbreviations

A&E: accident and emergency
ICU: intensive care unit
MLQ: Multifactor Leadership Questionnaire
ODQ: Organizational Description Questionnaire
SD: standard deviation
TAC: transactional culture
TAL: transactional leadership
TFC: transformational culture
TFL: transformational leadership

