## Supplementary material for "Organizational culture, quality of care and leadership style in government general hospitals in Kuwait: a multimethod study": Table 1

Table 1. Generic indicators information

| Indicator name | Indicator statement | Numerator statement | Denominator statement | Type of indicator | Data reported as | Target |
| --- | --- | --- | --- | --- | --- | --- |
| Discharge against medical advice | Percentage of patients discharged against medical advice in a hospital | The number of discharged against medical advice per department | Total number of hospital discharges per department excluding deaths or transfers to other hospitals | Outcome, rate-based | Percentage | Decrease in percentage |
| cancelled operations | Percentage of elective operations cancelled on the day of, or after admission | The number of last-minute cancelled operation lists | Total number of scheduled elective operations in the operation lists excluding A&E theater operations or day-case surgeries | Process, rate-based | Percentage | Decrease in percentage |
| Long post-appendectomy length of stay | Percentage of patients discharged from general surgical department after five days, or more from appendectomy | The number of patients discharged from general surgical department after five days, or more from appendectomy | Total number of patients discharged from general surgical department after five appendectomy | Outcome, rate-based | Percentage | Decrease in percentage |
| Non-operated discharges | Percentage of patients discharged from general surgical department without undergoing an operation | The number of patients discharged from general surgical department without undergoing an operation | Total number of patients discharged from the same surgical department excluding deaths or transfers to other hospitals | Process, rate-based | Percentage | Decrease in percentage |
| Unscheduled return for operations | The number of patients who have underwent a surgical procedure and returned to the operating theatre within the same hospital admission | Description of indicator population: Each patient who returned to the operating theater within the same admission | Not applicable | Outcome | Number | Decrease in number |
