## Supplementary material for "Organizational culture, quality of care and leadership style in government general hospitals in Kuwait: a multimethod study": Table 2

Table 2. Demographic and work-related characteristics of followers (respondents) in the six studied hospitals

| Demographic and work-related characteristic | Hospital (*n.* 271) | | | | | | Total  n (%) | *p* |
| --- | --- | --- | --- | --- | --- | --- | --- | --- |
|  | A  n (%) | B  n (%) | C  n (%) | D  n (%) | E  n (%) | F  n (%) |  |  |
| Age |  |  |  |  |  |  |  | .051 |
| 20 – 29 | 35 (12.9) | 45 (16.6) | 50 (18.5) | 39 (14.4) | 63 (23.2) | 40 (14.8) | 272 (16.7) |  |
| 30 – 39 | 120 (44.3) | 125 (46.1) | 129 (47.6) | 143 (52.8) | 118 (43.5) | 133 (49.1) | 768 (47.2) |  |
| 40 – 49 | 75 (27.7) | 62 (22.9) | 59 (21.8) | 66 (24.4) | 63 (23.2) | 60 (22.1) | 385 (23.7) |  |
| 50 + | 41 (15.1) | 39 (14.4) | 33 (12.2) | 23 (8.5) | 27 (10.0) | 38 (14.0) | 201 (12.4) |  |
| Gender |  |  |  |  |  |  |  | < .001 |
| Male | 87 (32.1) | 82 (30.3) | 106 (39.1) | 56 (20.7) | 101 (37.3) | 92 (33.9) | 524 (32.2) |  |
| Female | 184 (67.9) | 189 (69.7) | 165 (60.9) | 215 (79.3) | 170 (62.7) | 179 (66.1) | 1102 (67.8) |  |
| Nationality |  |  |  |  |  |  |  | < .001 |
| Kuwaiti | 27 (10.0) | 51 (18.8) | 25 (9.2) | 19 (7.0) | 48 (17.7) | 27 (10.0) | 197 (12.1) |  |
| Non-Kuwaiti | 244 (90.0) | 220 (81.2) | 246 (90.8) | 252 (93.0) | 223 (82.3) | 244 (90.0) | 1429 (87.9) |  |
| Education |  |  |  |  |  |  |  | .014 |
| Diploma | 104 (38.4) | 83 (30.6) | 101 (37.3) | 98 (36.2) | 94 (34.7) | 105 (38.7) | 585 (36.0) |  |
| Graduate | 104 (38.4) | 115 (42.4) | 101 (37.3) | 125 (46.1) | 87 (32.1) | 103 (38.0) | 635 (39.1) |  |
| Post-Graduate Diploma | 0 (0.0) | 1 (0.4) | 0 (0.0) | 0 (0.0) | 0 (0.0) | 1 (0.4) | 2 (0.1) |  |
| Masters | 36 (13.3) | 32 (11.8) | 39 (14.4) | 32 (11.8) | 44 (16.2) | 34 (12.5) | 217 (13.3) |  |
| PhD | 27 (10.0) | 40 (14.8) | 30 (11.1) | 16 (5.9) | 46 (17.0) | 28 (10.3) | 187 (11.5) |  |
| Current Position |  |  |  |  |  |  |  | .043 |
| Employee | 240 (88.6) | 242 (89.3) | 256 (94.5) | 251 (92.6) | 256 (94.5) | 249 (91.9) | 1494 (91.9) |  |
| Unit Head | 31 (11.4) | 29 (10.7) | 15 (5.5) | 20 (7.4) | 15 (5.5) | 22 (8.1) | 132 (8.1) |  |
| Period spent at current position |  |  |  |  |  |  |  | .012 |
| 1 – 9 | 181 (66.8) | 190 (70.1) | 195 (72.0) | 194 (71.6) | 211 (77.9) | 179 (66.1) | 1150 (70.7) |  |
| 10 – 19 | 56 (20.7) | 59 (21.8) | 60 (22.1) | 61 (22.5) | 41 (15.1) | 72 (26.6) | 349 (21.5) |  |
| 20 + | 34 (12.5) | 22 (8.1) | 16 (5.9) | 16 (5.9) | 19 (7.0) | 20 (7.4) | 127 (7.8) |  |
| Period spent at current hospital |  |  |  |  |  |  |  | .008 |
| 1 – 9 | 143 (52.8) | 157 (57.9) | 167 (61.6) | 179 (66.1) | 179 (66.1) | 164 (60.5) | 989 (60.8) |  |
| 10 – 19 | 78 (28.8) | 78 (28.8) | 75 (27.7) | 70 (25.8) | 59 (21.8) | 79 (29.2) | 439 (27.0) |  |
| 20 + | 50 (18.5) | 36 (13.3) | 29 (10.7) | 22 (8.1) | 33 (12.2) | 28 (10.3) | 198 (12.2) |  |
| Category |  |  |  |  |  |  |  |  |
| Physicians |  |  |  |  |  |  |  | .044 |
| Consultant | 6 (2.2) | 12 (4.4) | 10 (3.7) | 5 (1.8) | 4 (1.5) | 6 (2.2) | 43 (2.6) |  |
| Senior specialist | 3 (1.1) | 2 (0.7) | 6 (2.2) | 7 (2.6) | 8 (3.0) | 2 (0.7) | 28 (1.7) |  |
| Specialist/ Senior general practitioner A | 9 (3.3) | 8 (3.0) | 10 (3.7) | 4 (1.5) | 11 (4.1) | 5 (1.8) | 47 (2.9) |  |
| Senior registrar/ Senior general practitioner B | 10 (3.7) | 23 (8.5) | 15 (5.5) | 5 (1.8) | 25 (9.2) | 14 (5.2) | 92 (5.7) |  |
| Registrar/ General practitioner | 46 (17.0) | 33 (12.2) | 31 (11.4) | 30 (11.1) | 55 (20.3) | 41 (15.1) | 236 (14.5) |  |
| Nurses |  |  |  |  |  |  |  | < .001 |
| Head specialist | 5 (1.8) | 6 (2.2) | 1 (0.4) | 3 (1.1) | 2 (0.7) | 3 (1.1) | 20 (1.2) |  |
| Senior specialist | 9 (3.3) | 5 (1.8) | 7 (2.6) | 7 (2.6) | 4 (1.5) | 9 (3.3) | 41 (2.5) |  |
| Specialist | 24 (8.9) | 7 (2.6) | 11 (4.1) | 17 (6.3) | 12 (4.4) | 20 (7.4) | 91 (5.6) |  |
| Senior nurse | 73 (26.9) | 47 (17.3) | 64 (23.6) | 37 (13.7) | 43 (15.9) | 49 (18.1) | 313 (19.2) |  |
| Nurse | 81 (29.9) | 121 (44.6) | 112 (41.3) | 151 (55.7) | 103 (38.0) | 116 (42.8) | 684 (42.1) |  |
| Pharmacists |  |  |  |  |  |  |  | .225 |
| Head specialist | 0 (0.0) | 2 (0.7) | 1 (0.4) | 0 (0.0) | 1 (0.4) | 0 (0.0) | 4 (0.2) |  |
| Senior specialist | 2 (0.7) | 1 (0.4) | 0 (0.0) | 1 (0.4) | 0 (0.0) | 2 (0.7) | 6 (0.4) |  |
| Specialist | 0 (0.0) | 2 (0.7) | 0 (0.0) | 2 (0.7) | 0 (0.0) | 0 (0.0) | 4 (0.2) |  |
| Senior pharmacist | 3 (1.1) | 0 (0.0) | 1 (0.4) | 0 (0.0) | 2 (0.7) | 1 (0.4) | 7 (0.4) |  |
| Pharmacist | 0 (0.0) | 2 (0.7) | 2 (0.7) | 2 (0.7) | 1 (0.4) | 3 (1.1) | 10 (0.6) |  |

n: Number	(%): Percentage	*p*: *p*-value (Statistically significant at *p* ≤ .05, highly significant at *p* ≤ .001)
