## Supplementary material for "Organizational culture, quality of care and leadership style in government general hospitals in Kuwait: a multimethod study": Table 3

Table 3. Organizational culture styles based on the followers’ rating in departments of the six studied hospitals

| Department | Organizational Culture Type | Hospital | | | | | | | | | | | |  |  | *p^a^* |
| --- | --- | --- | --- | --- | --- | --- | --- | --- | --- | --- | --- | --- | --- | --- | --- | --- |
|  |  | A (n=271) | | B (n=271) | | C (n=271) | | D (n=271) | | E (n=271) | | F (n=271) | | Total (n=1626) | |  |
|  |  | n | (%) | n | (%) | n | (%) | n | (%) | n | (%) | n | (%) | n | (%) |  |
| Medicine | Transformational | 15 | (83.3) | 9 | (40.9) | 19 | (79.2) | 19 | (90.5) | 21 | (84.0) | 7 | (77.8) | 90 | (75.6) | .002 |
|  | Transactional | 3 | (16.7) | 13 | (59.1) | 5 | (20.8) | 2 | (9.5) | 4 | (16.0) | 2 | (22.2) | 29 | (24.4) |  |
| Surgery | Transformational | 8 | (61.5) | 11 | (57.9) | 6 | (66.7) | 6 | (85.7) | 8 | (47.1) | 7 | (53.8) | 46 | (59.0) | .636 |
|  | Transactional | 5 | (38.5) | 8 | (42.1) | 3 | (33.3) | 1 | (14.3) | 9 | (52.9) | 6 | (46.2) | 32 | (41.0) |  |
| Pediatrics | Transformational | 15 | (68.2) | 10 | (83.3) | 10 | (62.5) | 9 | (90.0) | 27 | (73.0) | 17 | (81.0) | 88 | (64.6) | .565 |
|  | Transactional | 7 | (31.8) | 2 | (16.7) | 6 | (37.5) | 1 | (10.0) | 10 | (27.0) | 4 | (19.0) | 30 | (25.4) |  |
| ICU | Transformational | 6 | (75.0) | 6 | (66.7) | 8 | (80.0) | 5 | (100.0) | 4 | (44.4) | 9 | (90.0) | 38 | (74.5) | .172 |
|  | Transactional | 2 | (25.0) | 3 | (33.3) | 2 | (20.0) | 0 | (0.0) | 5 | (55.6) | 1 | (10.0) | 13 | (25.5) |  |
| A&E | Transformational | 6 | (60.0) | 5 | (55.6) | 6 | (100.0) | 2 | (40.0) | 7 | (70.0) | 4 | (57.1) | 30 | (63.8) | .408 |
|  | Transactional | 4 | (40.0) | 4 | (44.4) | 0 | (0.0) | 3 | (60.0) | 3 | (30.0) | 3 | (42.9) | 17 | (36.2) |  |
| Laboratory | Transformational | 3 | (100.0) | 5 | (71.4) | 6 | (85.7) | 1 | (33.3) | 4 | (80.0) | 6 | (75.0) | 25 | (75.8) | .539 |
|  | Transactional | 0 | (0.0) | 2 | (28.6) | 1 | (14.3) | 2 | (66.7) | 1 | (20.0) | 2 | (25.0) | 8 | (24.2) |  |
| Nursing | Transformational | 141 | (73.4) | 130 | (69.9) | 160 | (82.1) | 189 | (87.9) | 126 | (76.8) | 150 | (76.1) | 896 | (78.0) | <.001 |
|  | Transactional | 51 | (26.6) | 56 | (30.1) | 35 | (17.9) | 26 | (12.1) | 38 | (23.2) | 47 | (23.9) | 253 | (22.0) |  |
| Pharmacy | Transformational | 3 | (60.0) | 4 | (57.1) | 2 | (50.0) | 5 | (100.0) | 1 | (25.0) | 4 | (66.7) | 19 | (61.3) | .352 |
|  | Transactional | 2 | (40.0) | 3 | (42.9) | 2 | (50.0) | 0 | (0.0) | 3 | (75.0) | 2 | (33.3) | 12 | (38.7) |  |
| Total | Transformational | 197 | (72.7) | 180 | (66.4) | 217 | (80.1) | 236 | (87.1) | 198 | (73.1) | 204 | (75.3) | 1232 | (75.8) | <.001 |
|  | Transactional | 74 | (27.3) | 91 | (33.6) | 54 | (19.9) | 35 | (12.9) | 73 | (26.9) | 67 | (24.7) | 394 | (24.2) |  |
| *p^b^* |  | .749 | | .163 | | .278 | | .016 | | .016 | | .508 | | .002 | |  |

n: Number	%: Percentage (per department)	ICU: Intensive Care Unit	A&E: Accident and Emergency

a: Difference between culture type in a department across all hospital (tested by Monte Carlo Exact Test)

b: Difference between culture type in a hospital across all departments (tested by Monte Carlo Exact Test)

*p*: *p*-value (Statistically significant at *p* ≤ .05, highly significant at *p* ≤ .001)
