## Supplementary material for "Organizational culture, quality of care and leadership style in government general hospitals in Kuwait: a multimethod study": Table 4

Table 4. Summary of leadership style, organizational culture style, and generic indicators at the six studied hospitals.

|  | Hospital | | | | | | Total | *p** |
| --- | --- | --- | --- | --- | --- | --- | --- | --- |
|  | A | B | C | D | E | F |  |  |
|  | n (%) | n (%) | n (%) | n (%) | n (%) | n (%) | n (%) |  |
| Leadership style |  |  |  |  |  |  |  | .674 |
| Transformational (TFL) | 228 (84.1) | 234 (86.3) | 233 (86.0) | 241 (88.9) | 229 (84.5) | 232 (85.6) | 1397 (85.9) |  |
| Transactional (TAL) | 43 (15.9) | 37 (13.7) | 38 (14.0) | 30 (11.1) | 42 (15.5) | 39 (14.4) | 229 (14.1) |  |
| Organizational culture style |  |  |  |  |  |  |  | < .001 |
| Transformational (TFC) | 197 (72.7) | 180 (66.4) | 217 (80.1) | 236 (87.1) | 198 (73.1) | 204 (75.3) | 1232 (75.8) |  |
| Transactional (TAC) | 74 (27.3) | 91 (33.6) | 54 (19.9) | 35 (12.9) | 73 (26.9) | 67 (24.7) | 394 (24.8) |  |
| Leadership style x Organizational culture style |  |  |  |  |  |  |  |  |
| TFL x TFC | 175 (64.6) | 164 (60.5) | 191 (70.5) | 218 (80.4) | 167 (61.6) | 180 (66.4) | 1095 (67.3) |  |
| TFL x TAC | 53 (19.6) | 70 (25.8) | 42 (15.5) | 23 (8.5) | 62 (22.9) | 52 (19.2) | 302 (18.6) |  |
| TAL x TFC | 22 (8.1) | 16 (5.9) | 26 (9.6) | 18 (6.6) | 31 (11.4) | 24 (8.9) | 137 (8.4) |  |
| TAL x TAC | 21 (7.7) | 21 (7.7) | 12 (4.4) | 12 (4.4) | 11 (4.1) | 15 (5.5) | 92 (5.7) |  |
| McNemar *p* | < .001 | < .001 | 0.068 | 0.533 | 0.002 | 0.002 | < .001 |  |
| Generic Indicators (2012) |  |  |  |  |  |  |  |  |
| % of discharge against medical advice | 5.0 | 1.0 | 4.9 | 7.6 | 2.2 | 7.9 |  | < .001 |
| % of cancelled operations | 11.3 | 12.9 | 9.5 | 9.3 | 12.3 | 10.4 |  | .047 |
| % of length of stay for appendectomy ≥ 5 days | 20.2 | 23.5 | 46.6 | 14.1 | 14.9 | 35.4 |  | < .001 |
| % of discharge without operation | 48.8 | 39.2 | 47.1 | 54.5 | 32.3 | 48.1 |  | < .001 |
| Unscheduled return for operations | 56 | 17 | 9 | 11 | 48 | 16 |  |  |

n: Number	%: Percentage	*: Monte Carlo Exact Test	TFL: Transformational leadership

TAL: Transactional leadership	TFC: Transformational culture	TAC: Transactional culture	*p*: *p*-value (Statistically significant at ≤ .05, highly significant at ≤ .001)
