## Supplementary material for "Organizational culture, quality of care and leadership style in government general hospitals in Kuwait: a multimethod study": Table 5

Table 5. Relationship between generic quality indicators and transformational leadership style of heads of departments in the six studied hospitals

| Generic quality indicators | Heads of departments transformational leadership style | |
| --- | --- | --- |
|  | r | *p* |
| Percentage of discharge against medical advice | -0.03 | .957 |
| Percentage of cancelled operations | -0.37 | .468 |
| Percentage of the length of stay for appendectomy ≥ 5 days | -0.09 | .872 |
| Percentage of discharge without operation | 0.49 | .329 |
| Number of unscheduled return for operations | -0.71 | .111 |

r: Pearson coefficient	*p*: *p*-value (Statistically significant at *p* ≤ .05, highly significant at *p* ≤ .001)

Correlation coefficient interpretation guidelines:^36^

>0.00 - 0.30: weak correlation	>0.30 – 0.70: moderate correlation	>0.70 – 1.00: strong correlation
