## Appendix 1 for "Organizational culture, quality of care and leadership style in government general hospitals in Kuwait: a multimethod study"

Appendix 1 – Mean scores of leadership styles according to followers’ rating (FR) and self-rating (SR) in all hospitals. The larger mean score of transformational (TFL) or transactional (TAL) factors indicates whether the leadership style is transformational or transactional.

| Hospitals | Leadership style | Director | | Medicine | | Surgery | | Pediatrics | | ICU | | A&E | | Laboratory | | Nursing | | Pharmacy | |
| --- | --- | --- | --- | --- | --- | --- | --- | --- | --- | --- | --- | --- | --- | --- | --- | --- | --- | --- | --- |
|  |  | FR  M ± SD | SR | FR  M ± SD | SR | FR  M ± SD | SR | FR  M ± SD | SR | FR  M ± SD | SR | FR  M ± SD | SR | FR  M ± SD | SR | FR  M ± SD | SR | FR  M ± SD | SR |
| A | TFL | 2.73 ± 1.09 | 2.75 | 3.27 ± 0.61 | 3.20 | 2.86 ± 0.81 | 3.65 | 2.45 ± 0.85 | 3.40 | 3.29 ± 0.39 | 3.55 | 3.03 ± 0.4 | 3.45 | 3.03 ± 0.5 | 3.60 | 2.79 ± 0.62 | 3.45 | 2.33 ± 0.6 | 3.8 |
|  | TAL | 2.16 ± 0.53 | 2.0 | 2.5 ± 0.5 | 2.67 | 2.31 ± 0.59 | 2.0 | 2.16 ± 0.45 | 2.58 | 2.57 ± 0.38 | 2.50 | 2.39 ± 0.46 | 2.17 | 2.58 ± 0.36 | 2.50 | 2.25 ± 0.50 | 2.25 | 2.52 ± 0.32 | 2.83 |
| B | TFL | 2.75 ± 0.82 | 3.35 | 2.51 ± 0.71 | 3.5 | 2.35 ± 0.62 | 3.6 | 2.60 ± 0.95 | 4.0 | 2.93 ± 0.69 | 2.40 | 2.23 ± 0.94 | 4.0 | 2.51 ± 1.10 | 3.05 | 2.67 ± 0.66 | 3.0 | 2.74 ± 0.53 | 3.45 |
|  | TAL | 2.44 ± 0.45 | 2.33 | 1.98 ± 0.34 | 2.58 | 2.06 ± 0.37 | 2.50 | 2.10 ± 0.48 | 2.83 | 2.31 ± 0.28 | 2.58 | 2.13 ± 0.41 | 2.67 | 2.0 ± 0.41 | 2.25 | 2.18 ± 0.45 | 2.42 | 2.21 ± 0.72 | 2.58 |
| C | TFL | 2.40 ± 0.94 | 3.75 | 2.93 ± 0.55 | 3.0 | 2.07 ± 1.0 | 3.50 | 2.95 ± 0.39 | 3.55 | 3.18 ± 0.48 | 3.55 | 3.34 ± 0.64 | 3.40 | 3.26 ± 0.47 | 3.15 | 2.60 ± 0.54 | 3.25 | 2.76 ± 0.58 | 3.45 |
|  | TAL | 2.07 ± 0.67 | 2.0 | 2.37 ± 0.51 | 1.75 | 2.23 ± 0.44 | 1.92 | 2.08 ± 0.35 | 2.25 | 2.55 ± 0.41 | 2.50 | 2.50 ± 0.39 | 2.33 | 2.36 ± 0.45 | 1.58 | 1.90 ± 0.14 | 3.0 | 2.19 ± 0.48 | 2.17 |
| D | TFL | 3.12 ± 0.29 | 3.45 | 2.96 ± 0.67 | 3.15 | 2.45 ± 0.52 | 3.55 | 3.32 ± 0.27 | 3.70 | 3.20 ± 0.58 | 3.55 | 2.49 ± 0.75 | 3.25 | 3.08 ± 0.25 | 3.80 | 2.80 ± 0.61 | 3.70 | 3.13 ± 0.3 | 3.50 |
|  | TAL | 2.37 ± 0.23 | 2.0 | 2.29 ± 0.52 | 2.25 | 2.17 ± 0.23 | 2.67 | 2.68 ± 0.44 | 2.08 | 2.15 ± 0.48 | 2.58 | 2.43 ± 0.26 | 2.33 | 1.78 ± 0.27 | 2.33 | 2.13 ± 0.44 | 2.58 | 2.25 ± 0.29 | 2.58 |
| E | TFL | 1.93 ± 0.67 | 3.0 | 3.01 ± 0.71 | 3.0 | 2.62 ± 0.7 | 3.50 | 2.65 ± 0.67 | 3.05 | 2.81 ± 0.55 | 3.75 | 2.79 ± 0.6 | 2.0 | 2.77 ± 0.83 | 3.80 | 2.64 ± 0.74 | 3.75 | 3.29 ± 0.44 | 3.80 |
|  | TAL | 1.86 ± 0.34 | 1.83 | 2.4 ± 0.46 | 1.83 | 2.14 ± 0.3 | 1.92 | 2.25 ± 0.56 | 1.75 | 2.15 ± 0.5 | 2.67 | 2.43 ± 0.52 | 2.0 | 2.55 ± 0.46 | 2.17 | 2.22 ± 0.64 | 2.17 | 3.0 ± 0.30 | 2.17 |
| F | TFL | 2.79 ± 0.81 | 3.35 | 3.46 ± 0.13 | 3.75 | 2.37 ± 0.86 | 3.90 | 3.09 ± 0.74 | 3.70 | 3.26 ± 0.44 | 3.50 | 2.72 ± 1.0 | 3.25 | 2.75 ± 0.36 | 2.0 | 2.60 ± 0.6 | 3.30 | 2.88 ± 0.53 | 3.45 |
|  | TAL | 2.39 ± 0.56 | 2.33 | 2.38 ± 0.19 | 1.42 | 2.19 ± 0.53 | 2.17 | 2.43 ± 0.59 | 2.33 | 2.46 ± 0.32 | 2.83 | 2.21 ± 0.62 | 2.25 | 2.04 ± 0.27 | 2.0 | 2.12 ± 0.41 | 2.67 | 2.24 ± 0.53 | 1.42 |

FR: Follower’s rating	SR: Self-rating	M: Mean	SD: Standard deviation

TFL: Transformational leadership	TAL: Transactional leadership	ICU: Intensive Care Unit	A&E: Accidents and Emergency
